# BNT162b2 boosted immune responses six months after heterologous or homologous ChAdOx1nCoV-19/BNT162b2 vaccination against COVID-19

**DOI:** 10.1101/2021.12.25.21268392

**Authors:** Georg M. N. Behrens, Joana Barros-Martins, Anne Cossmann, Gema Morillas Ramos, Metodi V. Stankov, Ivan Odak, Alexandra Dopfer-Jablonka, Laura Hetzel, Miriam Köhler, Gwendolyn Patzer, Christoph Binz, Christiane Ritter, Michaela Friedrichsen, Christian Schultze-Florey, Inga Ravens, Stefanie Willenzon, Anja Bubke, Jasmin Ristenpart, Anika Janssen, George Ssebyatika, Günter Bernhardt, Markus Hoffmann, Stefan Pöhlmann, Thomas Krey, Berislav Bošnjak, Swantje I. Hammerschmidt, Reinhold Förster

**Author notes:** Corresponding authors: Georg M.N. Behrens, Department of Rheumatology and Immunology, Hannover Medical School, Carl-Neuberg-Straße 1, D - 30625 Hannover, Germany, Reinhold Förster, Institute for Immunology, Hannover Medical School, Carl-Neuberg-Straße 1, D - 30625 Hannover, Germany. These authors contributed equally (Georg M. N. Behrens, Joana Barros-Martins, Anne Cossmann, Gema Morillas Ramos, Swantje I. Hammerschmidt, Reinhold Förster).

## Abstract

Reports suggest that COVID-19 vaccine effectiveness is decreasing, either due to waning immune protection, emergence of new variants of concern, or both. Heterologous prime/boost vaccination with a vector-based approach (ChAdOx-1nCov-19, ChAd) followed by an mRNA vaccine (e.g. BNT162b2, BNT) appeared to be superior in inducing protective immunity, and large scale second booster vaccination is ongoing. However, data comparing declining immunity after homologous and heterologous vaccination as well as effects of a third vaccine application after heterologous ChAd/BNT vaccination are lacking. We longitudinally monitored immunity in ChAd/ChAd (n=41) and ChAd/BNT (n=88) vaccinated individuals and assessed the impact of a second booster with BNT in both groups. The second booster greatly augmented waning anti-spike IgG but only moderately increased spike-specific CD4^+^ and CD8^+^ T cells in both groups to cell frequencies already present after the boost. More importantly, the second booster efficiently restored neutralizing antibody responses against Alpha, Beta, Gamma, and Delta, but neutralizing activity against B.1.1.529 (Omicron) stayed severely impaired. Our data suggest that inferior SARS-CoV-2 specific immune responses after homologous ChAd/ChAd vaccination can be cured by a heterologous BNT vaccination. However, prior heterologous ChAd/BNT vaccination provides no additional benefit for spike-specific T cell immunity or neutralizing Omicron after the second boost.

## Main text

While the COVID-19 vaccines currently approved by the European Medicine Agency (EMA) and U.S. Food and Drug Administration (FDA) provide high levels of protection against severe illness, the emergence of the Delta variant resulted in increasing numbers of breakthrough infections in fully vaccinated individuals (1). This coincided with evidence of waning immunity in vaccinated individuals (2, 3). Thus, booster vaccinations were proposed to reconstitute immunity and to potentially expand the breadth of immunity against SARS-CoV-2 variants of concern (VoC). Also policy makers have begun to promote booster vaccination not only for vulnerable patients but also to mitigate health-care and economic impact. Real world data confirm that booster vaccination is effective in preventing COVID-19 (4-6).

Concomitant to booster vaccination campaigns, the novel Omicron variant was recently identified in South Africa and its emergence was associated with a steep increase in cases and hospitalizations. Omicron is spreading rapidly in European, African and Asian countries as well as the USA, with case numbers doubling every two to three days (7). The S protein of the Omicron variant harbors an unusually high number of mutations, which increases immune evasion and potentially transmissibility (8-11). Thus, the Omicron variant constitutes a rapidly emerging threat to public health and might undermine global efforts to control the COVID-19 pandemic.

Heterologous prime boost strategies appear to offer immunological advantages to strengthen protection against COVID-19 achieved with currently available vaccines. Administration of mRNA vaccines like BNT162b2 (Comirnaty; BNT) after the initial ChAdOx1-nCov-19 (Vaxzevria, ChAd) dose as the second dose of a two-dose regimen was safe and had enhanced immunogenicity compared to homologous ChAdOx vaccination (12-18). We have previously reported on the results after homologous and heterologous vaccination after ChAd priming (16). Here we present findings from a subsequent analysis assessing the efficacy of a second booster vaccination after heterologous and homologous prime-boost vaccination on neutralization of VoC including Omicron.

In addition to our previously reported findings, we longitudinally monitored immunity after prime-boost COVID-19 vaccine treatment schedules and determined thereafter the impact of BNT booster (Methods). Health care professional vaccinees without previous SARS-CoV-2 infection, who had received ChAd/ChAd or ChAd/BNT, donated further blood four and six months after first booster vaccination and about two weeks after the second booster. The vaccination and blood collection schedule is depicted in Fig. 1A with additional demographic information (age and sex) in Extended Data Table 1. A third group of BNT/BNT vaccinees served as an independent control group for serologic analysis only and was monitored for up to nine months (Extended Data Table 1). As described (16), anti-SARS-CoV-2 spike IgG (anti-S IgG) were significantly higher in the ChAd/BNT group short after prime-boost vaccination when compared to the ChAd/ChAd group but declined significantly over time in both groups, with lower anti-S IgG after homologous vaccination prior to the second boost (Fig. 1B).

**Fig. 1.**
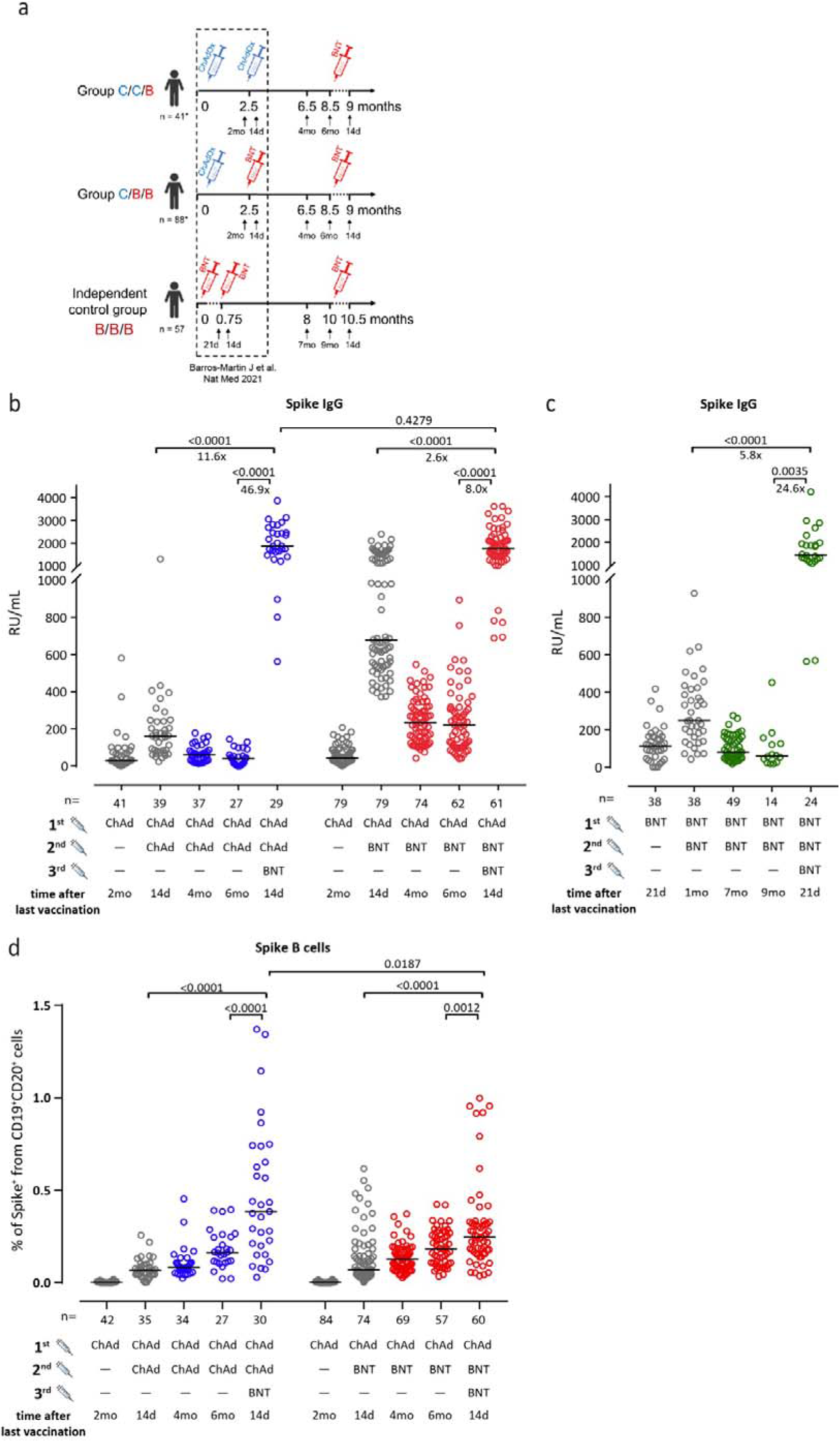
Participant recruitment schemes and humoral immune response. **a**, Participant recruitment and vaccination and blood sampling scheme. C, ChAd; B, BNT. **b**, A third heterologous or **c**, a third homologous immunization with BNT induce strong increases in anti S1 IgGs. Data are from **n** biologically independent samples as shown in the figure. **d**, A third heterologous immunization with BNT leads to increased frequencies of S-specific memory B cells. Data are from **n** biologically independent samples as shown in the figure. Statistics: **b**,**c**,**d**, Mixed effect analysis followed by Sidak’s multiple comparison test (within groups) and unpaired t test with Welch’s correction (between groups). The majority of the symbols depicted in grey had been published before (16, 26).

Following second booster immunization, we found greatly increased anti-S IgG responses in both groups. Boosting of the heterologous ChAd/BNT immunized group led to a significant 47.9 -fold increase for anti-S IgG (p<0.0001) and 8.0-fold increase in individuals after homologous ChAd vaccination (p<0.0001) (Fig. 1B). In both groups, anti-S IgG were considerably higher as compared to the first boost time point. More importantly, the second booster diminished previous differences between the heterologous and homologous prime-boost vaccination groups, since anti-S IgG were comparable in both groups after the second BNT boost and were within the range of triple BNT vaccinated individuals (Fig. 1C).

Next, we measured the frequency and phenotype of memory B cells carrying membrane-bound immunoglobulins specific for the Spike protein (Methods, Extended Data Fig. 1) over time. Interestingly, numbers of spike-specific memory B cells generated after prime-boost vaccination gradually increased during the following months with no significant difference between the ChAd/ChAd and the ChAd/BNT group (Fig. 1D). Again, the second booster with BNT led to a further and significant expansion of spike-specific memory B cells in both groups (Fig. 1D) in line with increased amounts of spike-specific antibodies, highlighting the impact of the second booster vaccination for better protection from SARS-CoV-2 infection.

For testing neutralizing activity of antibodies induced by vaccination, we employed our ELISA-based surrogate virus neutralization test (sVNT). We modified the sVNT to include Spike proteins of the Omicron variant (Methods) and for validation applied sera from vaccinees that had been recently tested for their neutralizing capacity applying vesicular stomatitis virus (VSV)-based pseudotyped virus neutralization assays (pVNT) (9, 20, 21). As for other VoC (16), we obtained a high degree of correlation between both assays with a R square value of 0.7044 (Extended Data Fig. 2). Thus, the sVNT is a reliable tool to quantitatively assess the neutralization capacity of vaccination-induced antibodies not only against the Wuhan but also against the Alpha, Beta, Gamma, Delta and Omicon variants of SARS-CoV-2.

Using the sVNT assays and consistent with declining anti-S IgG, we confirmed waning neutralizing activity against the Wuhan variant and particularly against VoC tested. Whilst the majority of participants had neutralizing antibodies against the Wuhan strain in pre-second boost plasma, neutralizing antibodies against the Alpha, Beta, Gamma and Delta variants were particularly in the CHAd/ChAd group less frequent or virtually absent (Fig. 2A). At 2 weeks after the second booster immunization, frequencies and titers of neutralizing antibodies against the Wuhan strain increased profoundly in the ChAd/BNT and ChAd/ChAd group with titers reaching values above those after the initial two injections in the latter group (Fig. 2A).

**Fig. 2.**
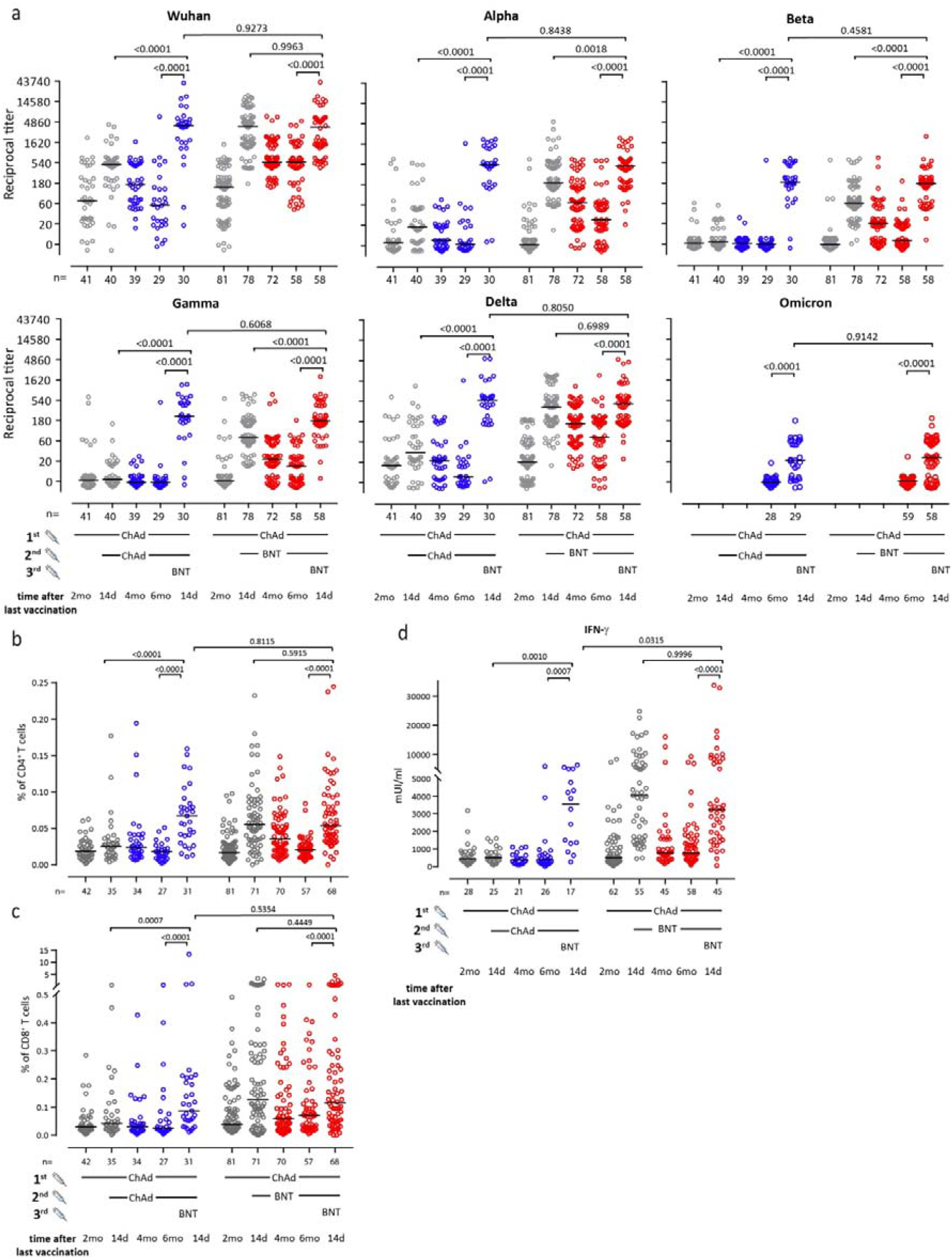
Heterologous vaccination induces neutralizing antibodies as well as CD4 and CD8 T cell responses. **a**, Heterologous ChAd/BNT/BNT or ChAd/ChAd/BNT vaccination induces neutralizing antibodies against Wuhan, B.1.1.7 (Alpha), B.1.351 (Beta), P.1 (B.1.1.28.1; Gamma), B.1.617.2 (Delta) and the B.1.1.529 (Omicron) SARS-CoV-2-S variants measured using the sVNT. Data are from **n** = biologically independent samples as indicated. For better visualization of identical titer values, data were randomly and proportionally adjusted closely around the precise titer results. Boost vaccination increased total percentage of cytokine-secreting CD4+ (**b**) and CD8+ (**c**) T cells. We calculated the total number of cytokine secreting cells as the sum of IFN-γ+TNF-α−, IFN-γ+TNF-α+ and IFN-γ−TNF-α+ cells in the gates indicated in Extended Data Fig. 4. Data are from **n** biologically independent samples as indicated. **d**, IFN-γ concentration in full blood supernatants after stimulation with SARS-CoV-2 S1 domain for 20–24 h measured in duplicate by IGRA (Euroimmun). Data are from **n** biologically independent samples as indicated. **a, b, c** and **d**. Mixed effect analysis followed by Sidak’s multiple comparison test (within groups) and unpaired t test with Welch’s correction (between groups). The majority of the symbols depicted in grey had been published before (16, 26).

Differences between the pre-booster vaccination regimens became even more evident when analyzing the neutralization capacity of antibodies induced against the VoC. In the ChAd/ChAd group, second booster immunization profoundly increased neutralization of the Alpha, Beta, Gamma and Delta variants (Fig. 2A), which was low for Alpha and Delta after prime/boost and virtually absent for Beta and Gamma. Whilst initial ChAd/BNT immunization had induced neutralizing antibodies at high levels against all analyzed VoC, the following decline was more than restored by the second booster (Fig. 2A). In fact, nearly all BNT boosted ChAd/BNT vaccinees had efficient neutralizing activity against Alpha, Beta, Gamma and Delta and amounts mostly above those from after the first booster. Importantly, the neutralization capacity against the Omicron variant was virtually absent before the second booster and remained low thereafter in comparison to the other VoC (Fig. 2A), with 9/29 (31%) and 27/58 (47%) of vaccinees in the ChAd/ChAd and ChAD/BNT group, respectively having no detectable neutralization activity after boost. We obtained very similar results after BNT booster in BNT/BNT vaccinated individuals (Extended Data Fig. 3). Altogether, these data indicate that the second booster immunization led to an increase of neutralizing antibodies in both vaccination groups against all tested VoC including the Omicron variant to only some extent.

Finally, we also analyzed frequencies and phenotypes of spike-specific T cells (Methods, Extended Data Fig. 4 and 5). We quantified numbers of spike-specific T cells as the sum of all cells producing IFN-γ or TNF-α as described previously (16). The frequencies of spike-specific CD4^+^ and CD8^+^ T cells in blood samples collected after the first booster vaccination were significantly higher in the ChAd/BNT group (Fig. 2B and C) and both cell populations declined over time, while they remained more or less stable after homologous vaccination. Whilst spike-specific CD4^+^ T cells declined to frequencies similar to individuals after homologous ChAd/ChAd vaccination (Fig. 2B), spike-specific CD8^+^ T cells remained above the frequencies of the heterologous vaccinated group (Fig. 2C). More interestingly, heterologous BNT booster in the ChAd/ChAd vaccination group significantly raised numbers of spike-specific CD4^+^ T cells above amounts observed after the first boost. In contrast, individuals with heterologous ChAd/BNT vaccination only regained spike-specific CD4^+^ T cell levels corresponding to levels after the first boost (Fig. 2B). Similarly, reboost with BNT did not result in an expansion of spike-specific CD8^+^ T cells above levels after first boost in ChAd/BNT vaccinees, but did so in ChAd/ChAd vaccinated individuals (Fig. 2C). Like for spike-specific CD4^+^ T cells, raised numbers in spike-specific IFN-γ-producing T cells in the ChAd/ChAd as well as the ChAd/BNT after BNT boost group was confirmed by cytokine measurement in supernatants after SARS-CoV-2 spike peptide stimulation (Fig. 2D). Again, BNT boost did not further increase spike-specific IFN-γ-producing T cells in ChAd/BNT vaccinated subjects above levels obtained already after first boost vaccination.

BNT booster potently increased anti-S IgG in all heterologous and homologous vaccinated individuals tested and this rise was accompanied by further strengthened neutralizing capacity against the Wuhan variant and Alpha, Beta, Gamma, and Delta. These data corroborate findings after homologous vaccination (22) and support current recommendations by the European Medicines Agency (EMA) and the European Centre for Disease Prevention and Control (ECDC). However, neutralization of the Omicron variant was absent before second booster and remained clearly inferior thereafter, irrespective of the previous vaccination scheme. Considering the kinetics of waning neutralizing antibodies against the other VoC after the first booster, we expect remaining neutralization against Omicron to vanish rapidly in the majority of vaccinees despite persisting high anti-S IgG concentrations. This suggests that variant-specific vaccines are required and need to be tested soon to better combat COVID-19 caused by Omicron and other emerging variants. In contrast, the second BNT booster only made up for absent rise in spike-specific T cell responses after homologous ChAd/ChAd vaccination and merely restored spike-specific CD4^+^ T cell responses after heterologous vaccination. These data confirm and extent reports that ChAd does not boost cellular responses after ChAd/ChAd vaccination (13, 23), but we are yet unable to make conclusion about the long-term protection and immunological memory. Although the relative role for T cell immunity remains unclear, spike-specific T memory cells are probably of great importance for protection against severe COVID-19, hospitalization and death. Our data reveal that additional BNT booster poorly support spike-specific CD8^+^ expansion and suggest that novel vaccines and vaccine schedules should be explored for further strengthening of adaptive cellular immunity against SARS-CoV-2 and its variants (24). Such vaccines should also aim to target other structural viral proteins including nucleocapsid and membrane proteins, which are less likely to be able to escape from capable immune recognition.

## Data Availability

All data produced in the present study are available upon reasonable request to the authors

## Acknowledgements

This work was supported by the German Center for Infection Research TTU 01.938 (grant no 80018019238 to G.M.N.B and R.F.), and TTU 04.820 to G.M.N.B., by Deutsche Forschungsgemeinschaft, (DFG, German Research Foundation) Excellence Strategy EXC 2155 “RESIST” (Project ID39087428 to R.F.), by funds of the State of Lower Saxony (14-76103-184 CORONA-11/20 to R.F.), by funds of the BMBF (NaFoUniMedCovid19 FKZ: 01KX2021; Projects B-FAST to R.F.) and Deutsche Forschungsgemeinschaft, SFB 900/3 (Projects B1, 158989968 to R.F.), and the European Regional Development Fund (Defeat Corona, ZW7-8515131 and ZW7-85151373 to G.M.N.B.). We thank the CoCo Study participants for their support and the entire CoCo study team for help. We would like to thank Luis Manthey, Annika Heidemann, Till Redeker, Madeleine Rommel, Christian Sturm, Marie Mikuteit, Jacqueline Niewolik, Ruth Sikora, Janine Topal, Kerstin Sträche, Birgit Heinisch, Michael Stephan, Mariel Nöhre, Simone Müller, Olivera Dragicevic, Kim Do Thi Hoang, Amy Kempf, and Inga Nehlmeier for technical and logistical support.

## Author Contributions Statement

Study design: G.M.N.B and R.F.

Data collection: J.B.-M., S.I.H., A.C., I.O., M.V.S., G.M.R., A.D.-J., L.H., M.K, G.P., C.B., C.R., M.F., C. S.-F., I.R., S.W., A.B., J.M., J.R., A.J., G.S., G.B., J.M., M.H., S.P., T.K.

Data analysis: G.M.N.B, J.B.-M., S.I.H., A.C., I.O., G.M.R., M.H., B.B, M.V.S.

Data interpretation: R.F., G.M.N.B.

Writing: G.M.N.B., R.F. with comments from all authors.

## Methods

### Participants

Participants for this analysis were from the COVID-19 Contact (CoCo) Study (German Clinical Trial Registry, DRKS00021152), an ongoing, prospective observational study monitoring anti-SARS-CoV-2 IgG immunoglobulin and immune responses in health care professionals (HCP) at Hannover Medical School and individuals with potential contact to SARS-CoV-2 (19, 25). An amendment from Dec 2020 allowed us to study the immune responses after COVID-19 vaccination. We followed the study cohort described previously (16) after heterologous ChAdOx/BNT or homologous ChAdOx/ ChAdOx and BNT/BNT vaccination. Scheduling appointments for a third booster vaccination with BNT was coordinated by an independent vaccination team according to vaccine availability.

One individual with previous SARS-CoV-2 infection as determined by positive anti-SARS-CoV-2 NCP IgG before vaccinations was excluded from this analysis. Two additional individuals of the ChAdOx/ ChAdOx group developed anti-SARS-CoV-2 NCP IgG after prime/boost vaccination and were excluded from follow up analysis. Demographics (sex and age) are depicted in Extended Data Table 1. After blood collection, we separated plasma from EDTA or lithium heparin blood (S-Monovette, Sarstedt) and stored it at −80 °C until use. We used full blood or isolated PBMCs from whole blood samples by Ficoll gradient centrifugation and for stimulation with SARS-CoV-2 peptide pools.

### Pseudotyped virus neutralization assay (pVNT)

pVNTs were performed at the Infection Biology Unit of the German Primate Center in Göttingen as described recently (21). Briefly, the rhabdoviral pseudotyped particles were produced in 293T cells transfected to express the desired SARS-CoV-2-S variant inoculated with VSV*DG-FLuc, a replication-deficient VSV vector that encodes for enhanced green fluorescent protein and firefly luciferase (FLuc) instead of VSV-G protein (kindly provided by Gert Zimmer, Institute of Virology and Immunology, Mittelhäusern, Switzerland). Produced pseudoparticles were collected, cleared from cellular debris by centrifugation and stored at −80 °C until used. For neutralization experiments, equal volumes of pseudotyped particles and heat-inactivated (56 °C, 30 min) plasma samples serially diluted in culture medium were mixed and incubated for 30 min at 37 °C. Afterwards, the samples together with non-plasma-exposed pseudotyped particles were used for transduction experiments. The assay was performed in 96-well plates in which Vero cells were inoculated with the respective pseudotyped particles/plasma mixtures. The transduction efficacy was analyzed at 16-18 hr post inoculation by measuring FLuc activity in lysed cells (Cell culture lysis reagent, Promega) using a commercial substrate (Beetle-Juice, PJK) and a plate luminometer (Hidex Sense Plate Reader, Hidex) with the Hidex Sense Microplate Reader Software (version 0.5.41.0).

### Serology

We measured SARS-CoV-2 IgG by quantitative ELISA (anti-SARS-CoV-2 S1 Spike protein domain/receptor binding domain IgG SARS-CoV-2-QuantiVac, Euroimmun, Lübeck, Germany) according to the manufacturer’s instructions (dilution up to 1:4000). We provide anti-S1 concentrations expressed as RU/mL as assessed from a calibration curve with values above 11 RU/mL defined as positive. These values can be converted in binding antibody units (BAU/mL) by multiplying RU/mL by 3.2. We performed anti SARS-CoV-2 nucleocapsid (NCP) IgG measurements according to the manufacturer’s instructions (Euroimmun, Lübeck, Germany). We used an AESKU.READER (AESKU.GROUP, Wendelsheim, Germany) and the Gen5 2.01 Software for analysis.

### Surrogate virus neutralization assay (sVNT) for SARS-CoV-2 variants

To determine neutralizing antibodies against the Wuhan-Spike, the B.1.1.7-Spike (Alpha), the B.1.351-Spike (Beta), the P.1-Spike (B.1.1.28.1; Gamma), the B.1.617.2 (Delta), and the B.1.1.529 (Omicron) variants of SARS-CoV-2-S in plasma, we modified our recently established surrogate virus neutralization test (sVNT) (20, 26). In this assay, the soluble receptor for SARS-CoV-2, ACE2, is bound to 96-well-plates to which different purified tagged receptor binding domains (RBDs) of the Spike-protein of SARS-CoV-2 can bind once added to the assay. Binding is further revealed by an anti-tag peroxidase-labelled antibody and colorimetric quantification. Pre-incubation of the Spike-protein with serum or plasma of convalescent patients or vaccinees prevents subsequent binding to ACE2 to various degrees, depending on the amount of neutralizing antibodies present. In detail, MaxiSorp 96F plates (Nunc) were coated with recombinant soluble hACE2-Fc(IgG1) protein at 300⍰ng per well in 50⍰μL coating buffer (30⍰mM Na2CO3, 70⍰mM NaHCO3, pH⍰9.6) at 4⍰°C overnight. After blocking with hACE2-Fc(IgG1), plates were washed with phosphate-buffered saline, 0.05% Tween-20 (PBST) and blocked with BD OptEIA Assay Diluent for 1.5 h at 37 °C. In the meantime, plasma samples were serially diluted threefold starting at 1:20 and then pre-incubated for 1⍰h at 37⍰°C with 1.5⍰ng recombinant SARS-CoV-2 Spike RBD of either the Wuhan strain (Trenzyme), the B.1.1.7 variant (N501Y; Alpha), the B.1.351 variant (K417N, E484K, N501Y; Beta) or the P.1 variant (K417T, E484K, N501Y; Gamma) (the latter three from SinoBiological), all with a C-terminal His-Tag. BD OptEIA Assay Diluent was used for preparing plasma sample as well as RBD dilutions. After pre-incubation with SARS-CoV-2 Spike RBDs, plasma samples were given onto the hACE2-coated MaxiSorp ELISA plates for 1 h at 37 °C. SARS-CoV-2 Spike RBDs pre-incubated with buffer only served as negative controls for inhibition. Plates were washed three times with PBST and incubated with an HRP-conjugated anti-His-tag antibody (clone HIS 3D5, provided by Helmholtz Zentrum München) for 1⍰h at 37⍰°C. Unbound antibody was removed by six washes with PBST. A colorimetric signal was developed on the enzymatic reaction of HRP with the chromogenic substrate 3,3’,5,5’-tetramethylbenzidine (BD OptEIA TMB Substrate Reagent Set). An equal volume of 0.2 M H_2_SO_4_ was added to stop the reaction, and the absorbance readings at 450⍰nm and 570⍰nm were acquired using a SpectraMax iD3 microplate reader (Molecular Devices) using SoftMAX Pro v7.03 software. For each well, the percent inhibition was calculated from optical density (OD) values after subtraction of background values as: Inhibition (%)⍰=⍰(1⍰−⍰Sample OD value/Average SARS-CoV-2 S RBD OD value)⍰×⍰100. Neutralizing sVNT titers were determined as the dilution with binding reduction⍰>⍰mean⍰+⍰2SD of values from a plasma pool consisting of three pre-pandemic plasma samples.

### SARS-CoV-2 protein peptide pools

We ordered 15 amino acid (aa) long and 10 aa overlapping peptide pools spanning the whole length of SARS-CoV2-Spike (-S) (total 253 peptides), -Membrane (-M) (43 peptides), -Nucleocapsid (-N) (82 peptides) or –Envelope (-E) (12 peptides; peptide no 4 could not be synthesized) proteins from GenScript. All lyophilized peptides were synthesized at >95% purity and reconstituted at a stock concentration of 50 mg/mL in DMSO (Sigma-Aldrich), except for 9 SARS-CoV2-S overlapping peptides (number 24, 190, 191, 225, 226, 234, 244, 245 and 246), 2 for SARS-CoV2-M (number 15 and 16), 1 for SARS-CoV2-N (number 61) and all 12 SARS-CoV2-E peptides that were dissolved at 25 mg/mL due to solubility issues. All peptides in DMSO stocks were stored at −80 °C until used.

### T cell re-stimulation assay

PBMCs, isolated using a Ficoll gradient, were re-suspended at concentration of 20 × 10^6^ cells/mL in complete RPMI medium [RPMI 1640 (Gibco) supplemented with 10% FBS (GE Healthcare Life Sciences, Logan, UT), 1mM sodium pyruvate, 50 µM β-mercaptoethanol, 1% streptomycin/penicillin (all Gibco)]. For stimulation, cells were diluted with equal volume of peptide pools containing S-protein or mixture of M-, N- and E-proteins. Peptide pools were prepared in complete RPMI containing brefeldin A (Sigma-Aldrich) at final concentration of 10 µg/mL. In the final mixture each peptide had concentration of 2 µg (∼1.2 nmol)/mL, except for SARS-CoV2-S peptides number 24, 190, 191, 225, 226, 234, 244, 245 and 246, SARS-CoV2-M peptides 15 and 16, and SARS-CoV2-N peptide 61, which were used at final concentration of 1 µg/mL due to solubility issues. As a negative control, we stimulated the cells with DMSO, used in maximal volume corresponding to DMSO amount in peptide pools (equaling to 5 % DMSO in final medium volume) Extended Fig. 5. In each experiment, we used cells stimulated with Phorbol-12-myristate-13-acetate (PMA; Calbiochem) and ionomycin (Invitrogen) at final concentration of 50 ng/mL and 1500 ng/mL, respectively, as an internal positive control. Cells were then incubated for 12-16 hr at 37 °C, 5% CO2. After washing, cells were resuspended in MACS buffer (PBS supplemented with 3% FBS and 2mM EDTA). Non-specific antibody binding was blocked by incubating samples with 10% mouse serum at 4 °C for 15 min. Next, without washing, an antibody mix of anti-CD3-AF532 (UCHT1; #58-0038-42; Lot # 2288218; Invitrogen; 1:50), anti-CD4-BUV563 (RPA-T4; #741353; Lot # 9333607; BD Biosciences; 1:200), anti-CD8-SparkBlue 550 (SK1; #344760; Lot #B326454; Biolegend; 1:200), anti-CD45RA (HI100, #740298, Lot # 0295003; BD Biosciences; 1:200), anti-CCR7 (G043H7; #353230; Lot # B335328; Biolegend; 1:50), anti-CD38 PerCP-eF710 (HB7; #46-0388-42; Lot # 2044748; Invitrogen; 1:100) and Zombie NIR(tm) Fixable Viability Kit (#423106; Lot # B323372; BioLegend) was added. After staining for 20 min at RT, cells were washed before they were fixed and permeabilized (#554714; BD Biosciences) according to the manufacturers’ protocol. Next, intracellular cytokines were stained using anti-IFN-PE-Cy7 (B27; #506518; Lot # B326674; Biolegend; 1:100), anti-TNF-AF700 (Mab11; #502928; Lot # B326186; Biolegend; 1:50) and anti-IL-17A-BV421 (BL168; #512322; Lot # B317903; Biolegend; 1:50) for 45 min on RT. Excess antibodies were washed away and cells were then acquired on Cytek Aurora spectral flow cytometer (Cytek) equipped with five lasers operating on 355nm, 405nm, 488nm, 561nm and 640nm (for gating strategy, see Extended Data Fig. 4). All flow cytometry data was acquired using SpectroFlo v2.2.0 (Cytek) and analyzed FCS Express V7 (Denovo).

### Flow cytometric analysis of Spike-specific B cells

Total leukocytes were isolated from whole blood using erythrolysis in 0.83% ammonium chloride solution. Isolated cells were then washed, counted and resuspended in PBS and stained for 20 min on RT with an antibody mix containing antibodies listed in Extended Data Figure 1A together with Spike-mNEONGreen protein (5μg per reaction; production will be described elsewhere). After one wash, samples were acquired on spectral flow cytometer and the data was analyzed as described above (for gating strategy, see Extended Data Fig. 1B).

### Quantification of IFN-γ release

0.5 mL full blood were stimulated with manufacturer’s selected parts of the SARS-CoV-2 S1 domain of the Spike Protein for a period of 20-24 h. We carried out negative and positive controls according to the manufacturer’s instruction and measured IFN-*γ* using an ELISA (SARS-CoV-2 Interferon Gamma Release Assay, IGRA (Euroimmun, Lübeck, Germany). For analysis, we used an AESKU.READER (AESKU.GROUP, Wendelsheim, Germany) and the Gen5 2.01 Software.

### Statistics

Statistical analysis was done using GraphPad Prism 8.4 (GraphPad Software, USA) and SPSS 20.0.0 (IBM SPSS Statistics, USA). For comparison of levels of Spike-specific IgG levels, as well as for comparison of percentages of cytokine-secreting T cells, for comparision of frequencies of Spike-specific B cells, or cytokine concentrations in the IGRA assay and sVNT values we used mixed effect analysis with Sidiak’s multiple comparison paired t-test (within groups) or unpaired T test with Welch correction 2-way ANOVA followed by Sidak’s multiple comparison test (between groups). Percentages of cytokine secreting T cells were log transformed prior comparison. For comparison of sVNT titers we used Chi-square test for trend. Differences were considered significant if p < 0.05. Correlation between sVNT and pVNT values was calculated using single linear regression analysis.

### Ethics committee approval

The CoCo Study and the analysis conducted for this article were approved by the Internal Review Board of Hannover Medical School (institutional review board no. 8973_BO-K_2020, amendment Dec 2020).

## Data availability statement

All requests for raw and analyzed data that underlie the results reported in this article will be reviewed by the CoCo Study Team, Hannover Medical School to determine whether the request is subject to confidentiality and data protection obligations. Data that can be shared will be released via a material transfer agreement.

## Extended Data

**Extended Data Table. 1.**
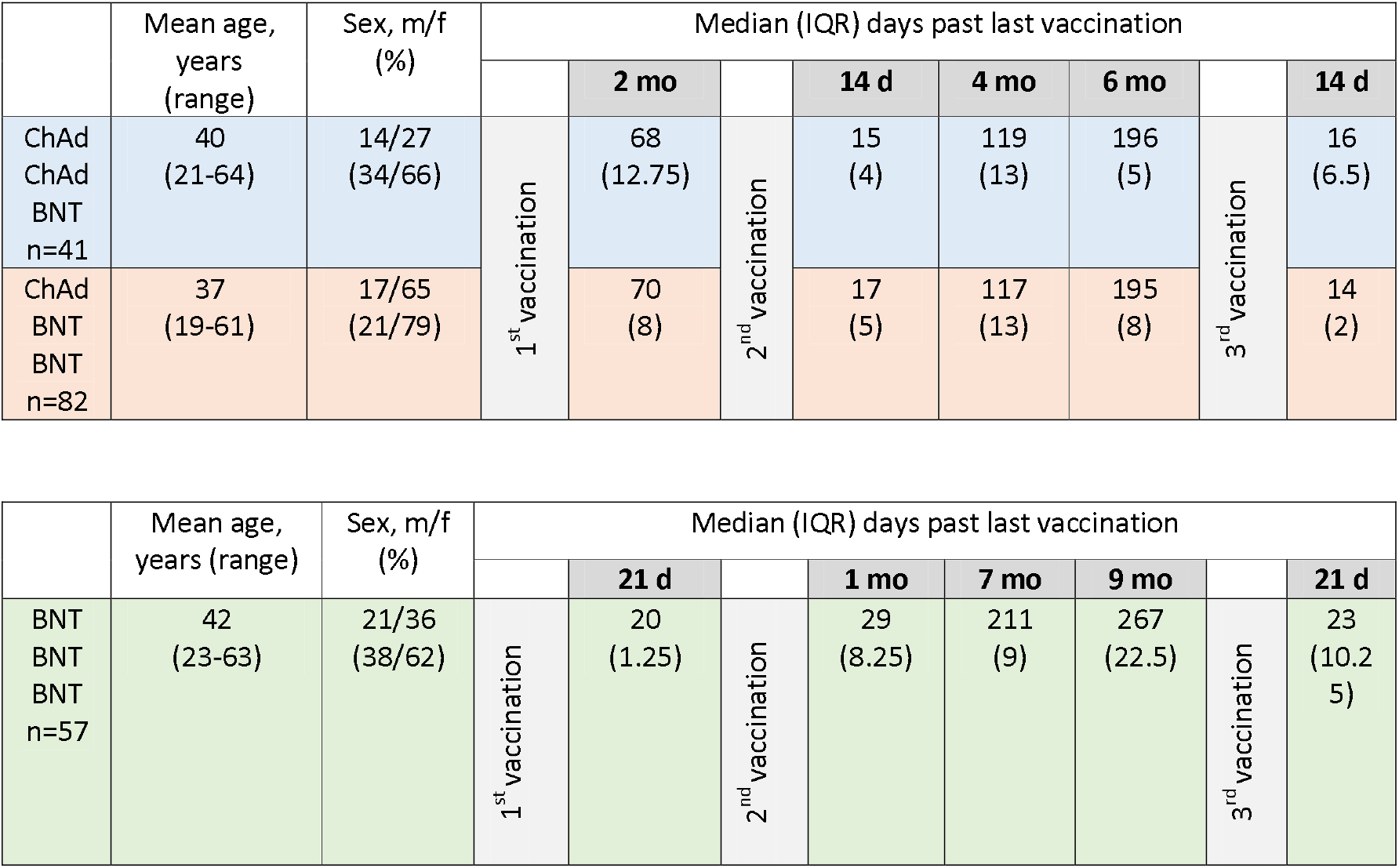
Demographic data and median time in days since last vaccination for the five blood collection time points (months = mo and days = d) of the three vaccination groups.

**Extended Data Fig. 1.**
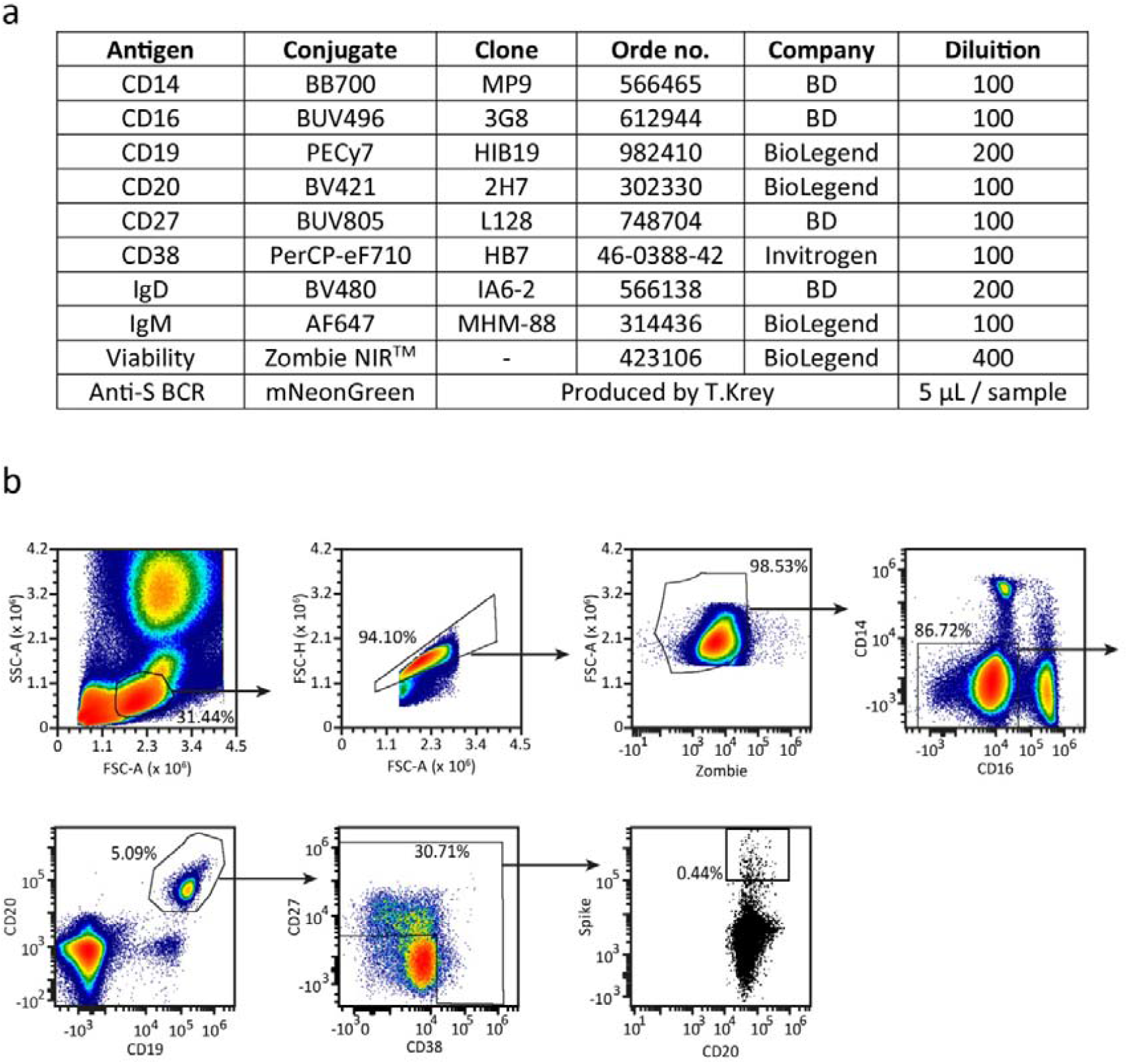
Antibody panel **a** and gating strategy **b** for SARS-CoV-2-S (Spike)-specific B cell populations in blood. Pseudocolor plots show representative data from a female donor 283 days after priming with ChAd; 213 days after a second dose with BNT and 14 days after a third dose with BNT.

**Extended Data Fig. 2.**
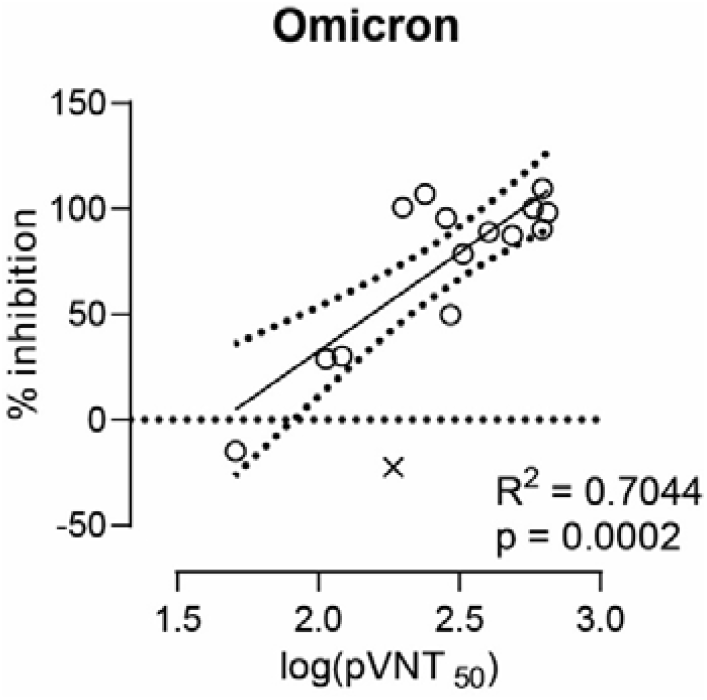
Antibody neutralization measurements against the Omicron SARS-CoV-2 variant is positively correlated between the virus neutralization tests (sVNT) and pseudotyped virus neutralization tests (pVNT). Correlation (solid line) and 95% confidence intervals (dotted lines) between sVNT1:20 and antibody titers resulting in 50% reduction of luciferase activity in pVNT, indicated as pVNT50. Open circles, values from individual donors, outliers are marked with X and were defined as values with absolute residual value > 2 SD of all residual values in each group of samples. Correlation was calculated using single linear regression.

**Extended Data Fig. 3.**
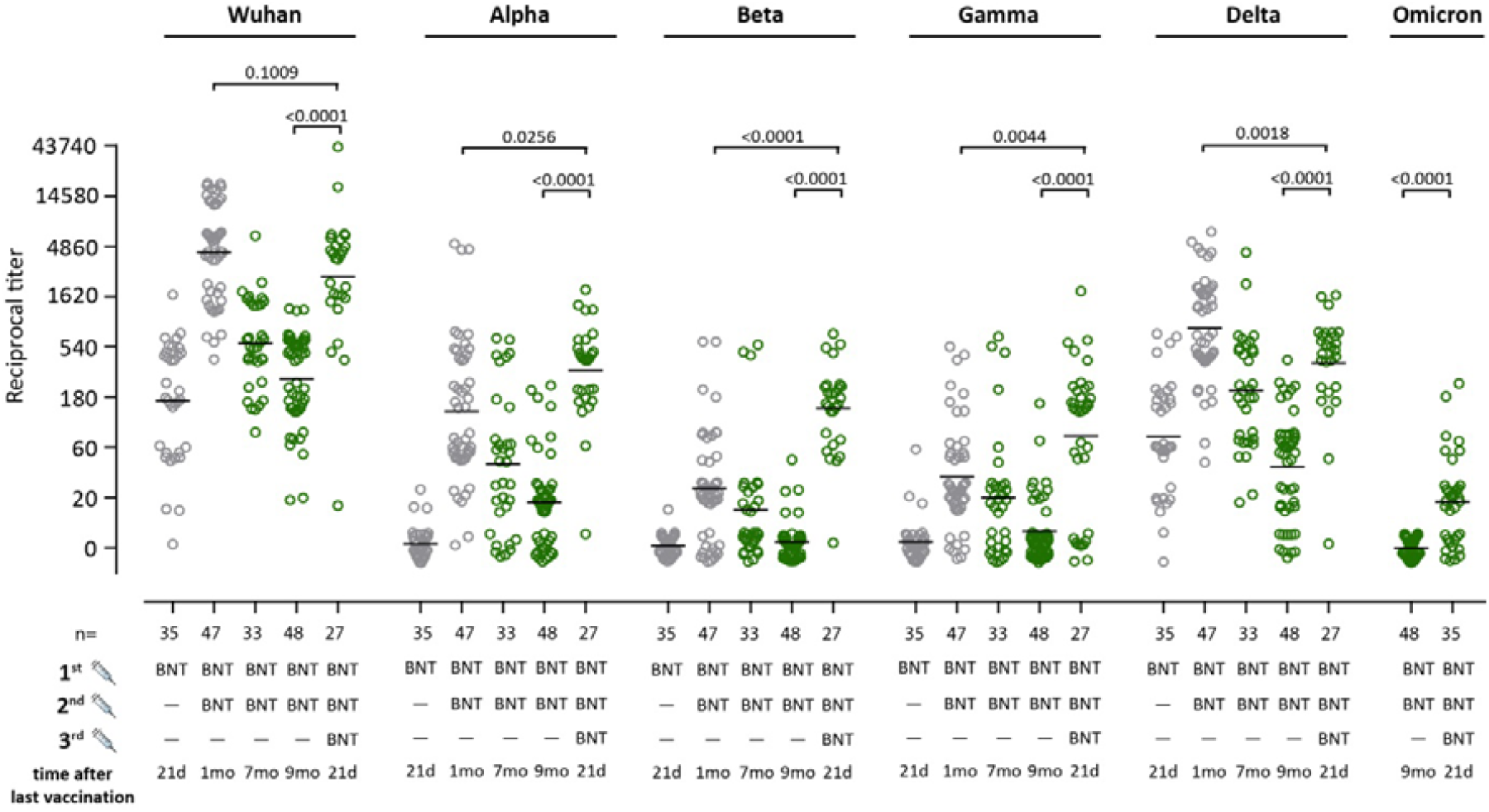
Humoral immune response against all SARS-CoV-2 variants following homologous BNT162b2 (BNT) / BNT /BNT vaccination. Reciprocal titers of neutralizing antibodies against Wuhan, B.1.1.7 (Alpha), B.1.351 (Beta), P.1 (B.1.1.28.1; Gamma), B.1.617.2 (Delta) and the B.1.1.529 (Omicron) SARS-CoV-2-S variants measured using the sVNT. Data are from *n* = biologically independent samples as indicated. Mixed effect analysis followed by Sidak’s multiple comparison test (within groups). For better visualization of identical titer values, data were randomly and proportionally adjusted closely around the precise titer results. The majority of the symbols depicted in grey had been published before (16, 26).

**Extended Data Fig. 4.**
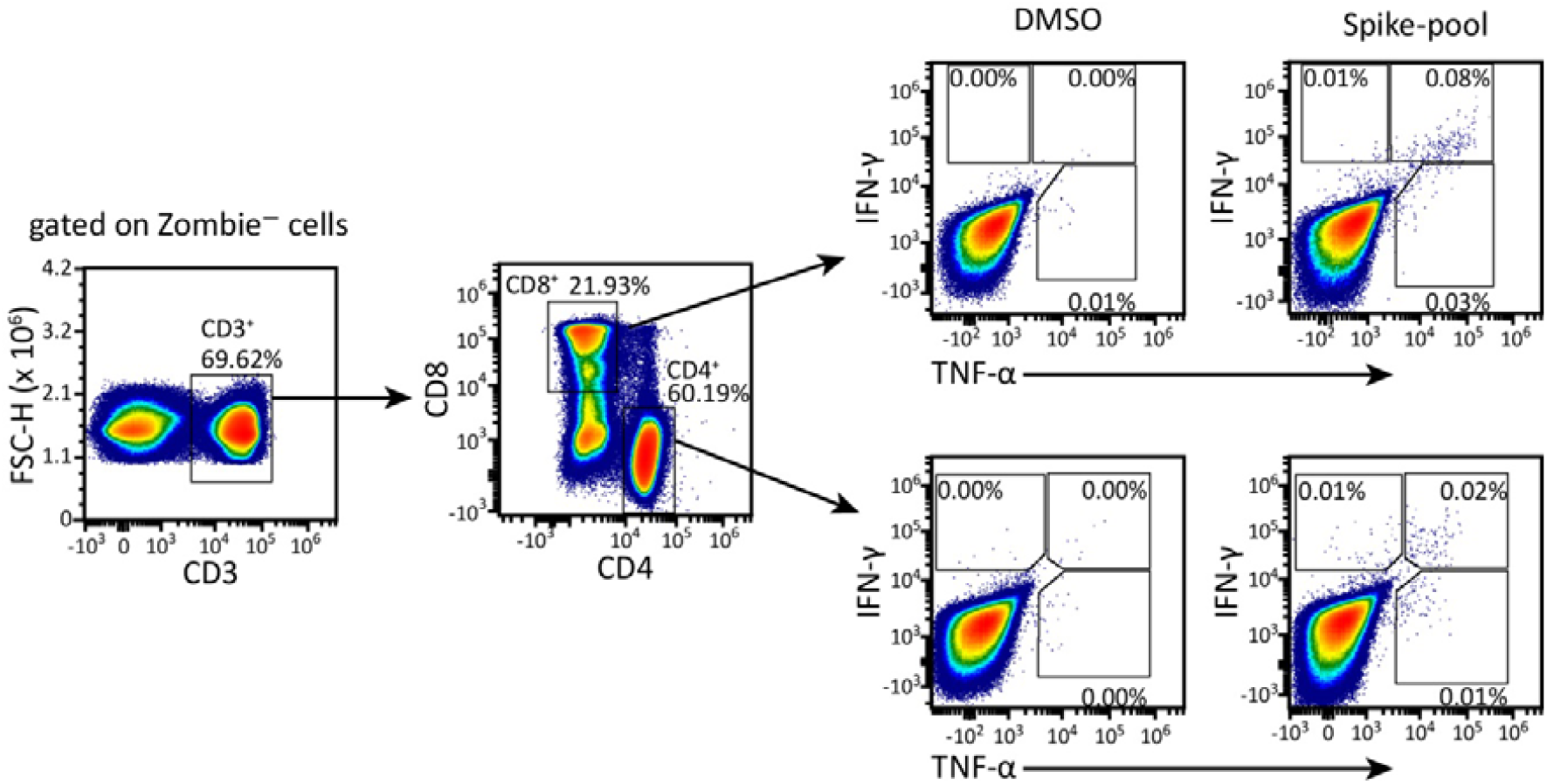
Gating strategy used for detection of cytokine producing CD4+ and CD8+ T cells after *ex vivo* re-stimulation with DMSO or the pool of Spike-specific peptides for 12–16 hr.

**Extended Data Fig. 5.**
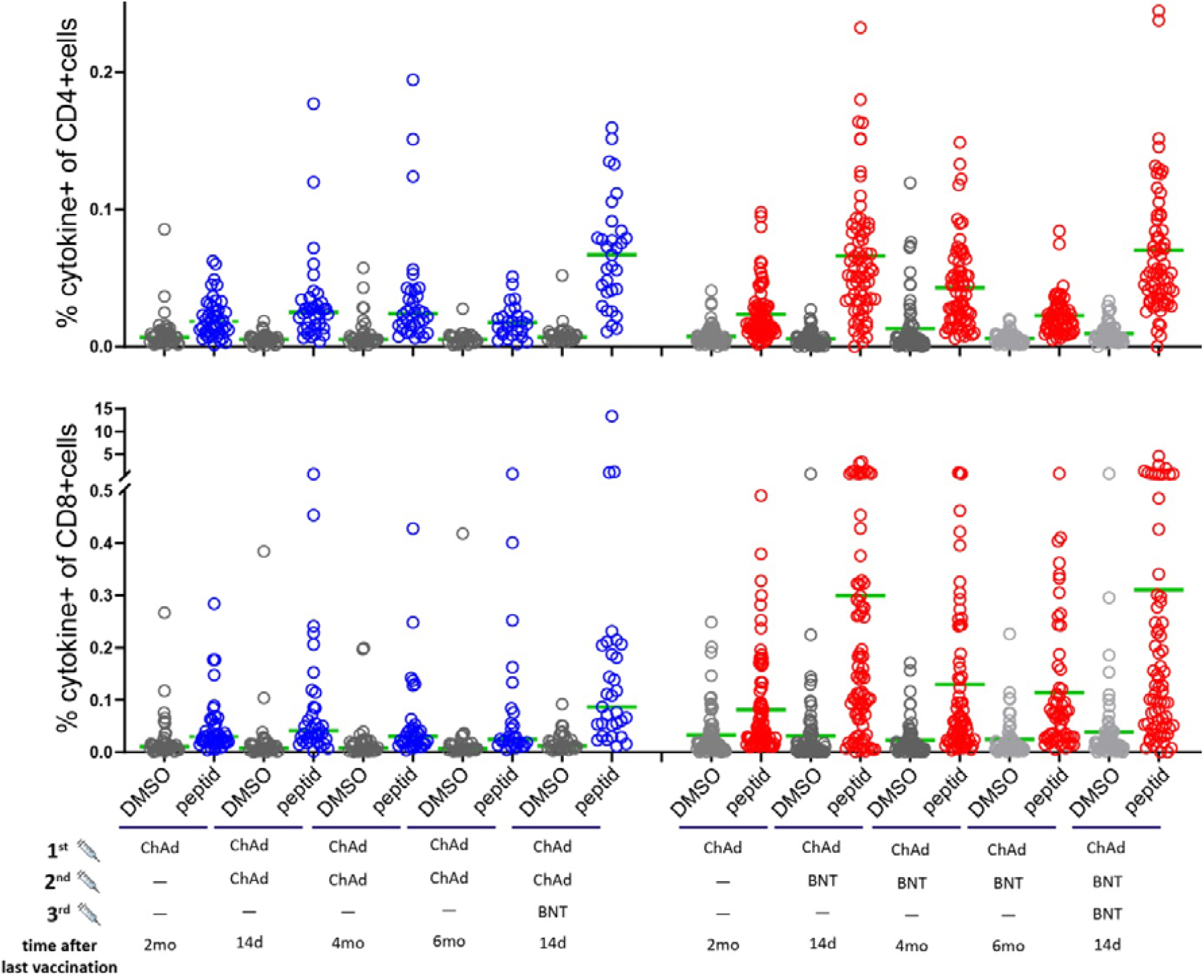
Frequency of cytokine-producing CD4^+^ T cells and CD8^+^ T cells after *ex vivo* re-stimulation with DMSO or the pool of Spike-specific peptides for 12–16 hr.

